# Impact of the COVID-19 pandemic on the frequency of donations and transfusions in a Peruvian blood bank

**DOI:** 10.1101/2025.08.11.25333474

**Authors:** Angelo De-La-Torre-Ugarte, Miryam Danitza More Yupanqui, Abraham De-Los-Rios-Pinto, Isabel Pinedo Torres

**Author notes:** All authors contributed substantially to this work. Funding: The study was self-funded by the authors. Competing interests: The authors have declared that no competing interests exist.

## Abstract

**Background:** The COVID-19 pandemic disrupted blood donation and transfusion services globally. In Peru, where the system relies heavily on replacement donors, this crisis exposed structural weaknesses and affected blood availability. Therefore, the aim of this study was to determine the impact of the COVID-19 pandemic on the frequency of blood donations and transfusions in a Peruvian blood bank.

**Methods:** A retrospective analytical study was conducted based on the review of secondary databases from the blood bank. Monthly averages of blood donations and transfusions were compared between a prepandemic and a pandemic period. Differences were calculated and statistical significance was assessed.

**Results:** A total of 14 058 donation records and 21 747 transfusion records were analyzed. Total donations decreased by 44.15% with the greatest reduction observed among repeat donor (–56.43%). Voluntary donations increased by 13.79%, but remained below 5% of the total. Transfusions declined by 38.5%, with reductions observed across all blood components. Hospital transfusion requests also decreased (–40.11%), especially in the surgical area (– 59.22%). Inferential analysis showed statistically significant reductions in monthly donations (mean difference: 256.75; 95% CI: 167.46 to 346.04; p < 0.01) and monthly transfusions (mean difference: 324.85; 95% CI: 213.35 to 436.35; p < 0.01).

**Conclusions:** The COVID-19 pandemic exposed the fragility of the Peruvian transfusion system, significantly reducing blood availability. Strengthening voluntary donor recruitment strategies, improving inventory management, and developing contingency plans are essential to ensure blood supply during future health crises.

## Introduction

Blood banks are a cornerstone of healthcare systems, providing essential support for surgeries, emergencies, and chronic treatments [1,2]. heir operation relies primarily on voluntary donors [3,4]. However, adverse events such as natural disasters, conflicts, and pandemics disrupt the blood supply chain, causing fluctuations in supply and demand that reveal the system’s volatility and underscore the need for efficient strategies to maintain its stability [1,3,5,6].

The COVID-19 pandemic significantly impacted blood donation and transfusion services [1,2]. Countries including China, Spain, India, the United States, Canada, and Peru reported donation declines ranging from 20% to 70%, mainly due to mobility restrictions, fear of contagion, the suspension of donation campaigns, and reduced elective surgeries, which also decreased demand [4,7–9]. This crisis affected donor willingness and introduced new challenges in ensuring donor safety and screening, especially given the potential risk of viral transmission [4,10,11].

The magnitude and characteristics of the pandemic’s impact varied depending on socioeconomic, organizational, and structural factors unique to each healthcare system [3,5]. To mitigate shortages, countries adopted digital campaigns, mobile donation units, and rational inventory management policies [1,6]. Retaining regular donors also emerged as a critical strategy for strengthening blood supply resilience [4,6]. The pandemic underscored the importance of dynamic planning to manage fluctuating donation rates and transfusion demands, preventing both shortages and wastage of blood components [3,5].

In Latin America and particularly in Peru these challenges are exacerbated by fragmented health systems, limited budgets, and weak interinstitutional coordination [4,7]. During the pandemic, Peru reported up to a 70% drop in blood donations, severely compromising transfusion-dependent care [12]. Specific causes of donor deferral and shifts in donor demographics were also noted, highlighting the need for context-based research to develop strategies ensuring blood safety and supply sustainability during future crises [4,7].

This study aims to analyze the impact of the COVID-19 pandemic on the frequency of blood donations and transfusions at a Peruvian blood bank. The findings intend to inform the development of effective policies and strategies for blood supply management in critical health contexts.

## Materials and Methods

### Studyd design and context

An observational, retrospective, analytical study was conducted based on the analysis of a secondary database from the Blood Bank of the Hospital Nacional Daniel Alcides Carrión. This high-complexity hospital serves an estimated population of 995,494 inhabitants and is located in the district of Bellavista, Callao, Peru. The institution is classified as a Type II hemotherapy center, as it performs donor selection and training activities.

### Population and sample

Access to the digital database was granted on November 1, 2023, for data collection and subsequent analysis in two groups: hospitalized patients for whom blood transfusions were requested, and donors registered in the hospital’s Blood Bank. All completed transfusion requests and blood donations were included (census approach), as recorded in the institution’s digital database, corresponding to adult patients aged 18 to 60 years, the upper age limit for blood donation in Peru during the period from January 1, 2019, to March 6, 2021.

This period was subdivided into two phases for comparative analysis: a prepandemic year, from January 1, 2019, to March 5, 2020, and a pandemic year including the first wave and part of the second wave of COVID-19 in Peru from March 6, 2020 (the date of the first confirmed case of COVID-19 in the country) to March 6, 2021, as reported by the Peruvian national health system. Records with incomplete information were excluded, particularly those lacking data on blood group.

### Variable operationalization

The dataset included *variables related to blood donation*, such as: monthly donation frequency (number of donations per month), average donor age (mean donor age per month), number of male donors per month, number of new donors (individuals donating for the first time), number of repeat donors (individuals with at least one previous donation event), number of voluntary donors per month, and number of replacement donors per month (individuals who replaced blood units received by friends or relatives during hospitalization).

Regarding donation characteristics, the following monthly indicators were evaluated: total number of donated blood units and ABO blood group distribution (total number of donations by blood type: A+, A−, B+, B−, AB+, AB−, O+, O−, respectively).

*Variables related to transfusion* that were evaluated on a monthly basis included: total number of transfusions, number of whole blood units transfused, number of platelet units transfused, number of cryoprecipitate units transfused, total number of fresh frozen plasma units transfused, and the number of transfusion requests originating from critical care units, surgical wards, and general hospitalization areas (referring to requests for any blood component).

### Data analysis

To evaluate the impact of the COVID-19 pandemic on the Blood Bank, a comparative analysis was conducted between the two defined periods: the pre-pandemic period and the pandemic period. Monthly averages of blood donations and transfusions were calculated for each period, and the mean differences between the two periods were estimated to quantify the changes observed during the pandemic. These differences were reported along with their 95% confidence intervals and corresponding p-values to assess statistical significance.

The statistical power of the study was calculated using the monthly mean and standard deviation (SD) of the number of transfusions during both periods: pre-pandemic (mean: 961.93; SD: 132.45) and pandemic (mean: 637.08; SD: 148.84). The analysis yielded a power of 100%, ensuring the validity of the comparative evaluation.

The statistical analysis was performed using Stata version 16. Quantitative variables were described using means or medians, depending on data distribution, while categorical variables were summarized as frequencies and percentages. For bivariate analysis, Student’s t-test or the Mann–Whitney U test was used, depending on whether the assumptions of normality and homogeneity of variances were met. A p-value < 0.05 was considered statistically significant, with a 95% confidence interval.

### Ethical considerations

This was a non interventional study based on secondary database that initially contained identifiable participant information. All data were treated with strict confidentiality and completely anonymized before any analysis was performed. It was approved by the Ethics Committee of Universidad Científica del Sur under registration number 804-2021-PRE15, and it received the corresponding institutional authorization (Document No. 3350-2023-HNDAC) for access to the Blood Bank database.

## Results

A total of 37,662 electronic records corresponding to blood donation and transfusion events were identified. Among these, 15,915 records referred to blood donations, of which 14,058 valid records remained after applying the selection criteria. These were distributed as follows: 9,020 records during the pre-pandemic period and 5,038 records during the pandemic period. In addition, 21,747 records corresponding to transfusions were identified, all of which were included in the analysis. Of these, 13,467 belonged to the pre-pandemic period and 8,280 to the pandemic period (Fig 1).

**Fig 1.**
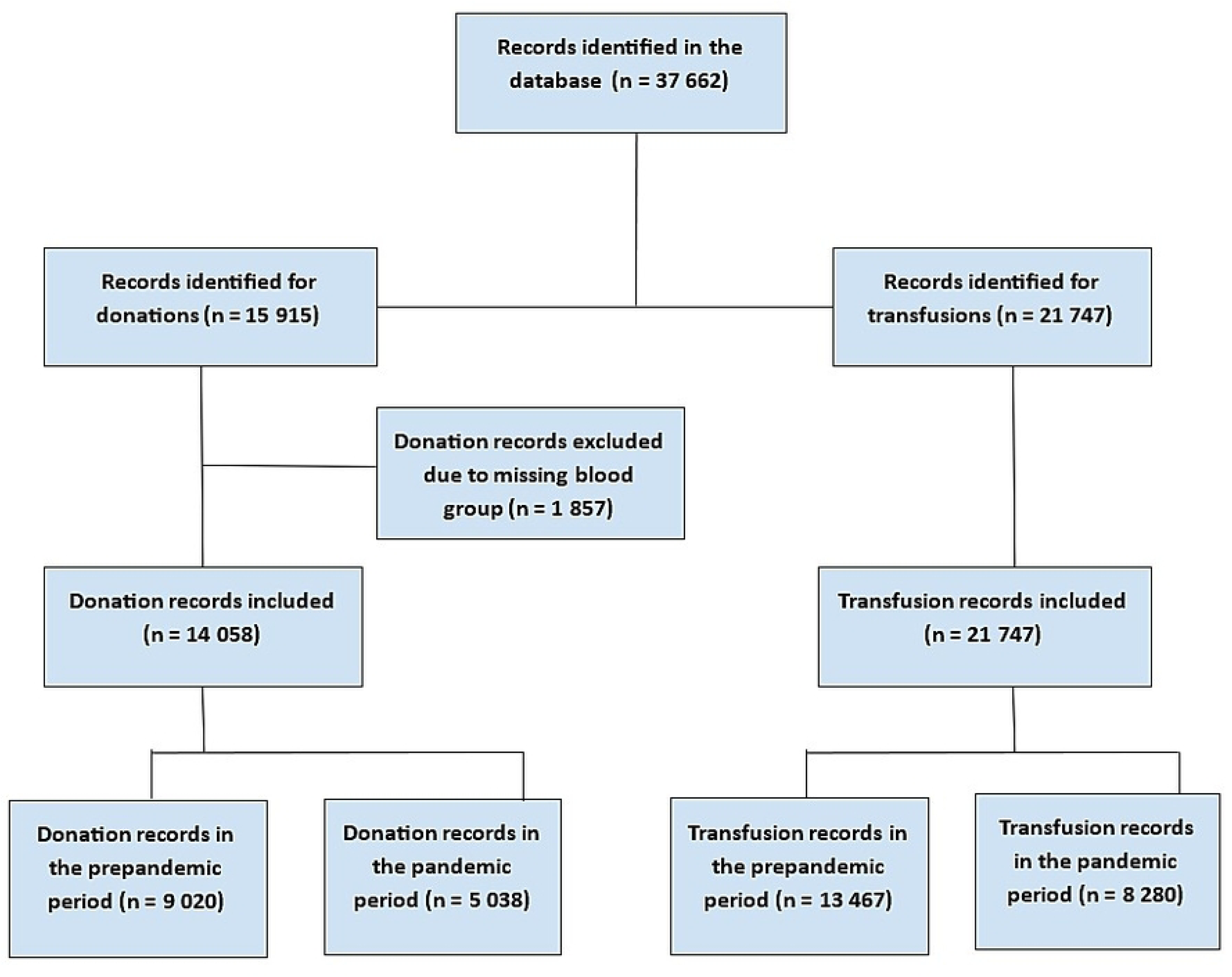
Flow diagram of blood donation and transfusion records included in the study during the analyzed period A 44.15% reduction in the total number of donations was observed. This decrease primarily affected replacement donors, who declined by 45.31%, and repeat donors, who showed a 56.43% drop. Additionally, the number of new donors decreased by 31.64%. Although there was a 13.79% increase in voluntary donations, their proportion relative to the total remained below 5% in both periods (Fig 2).

**Fig 2.**
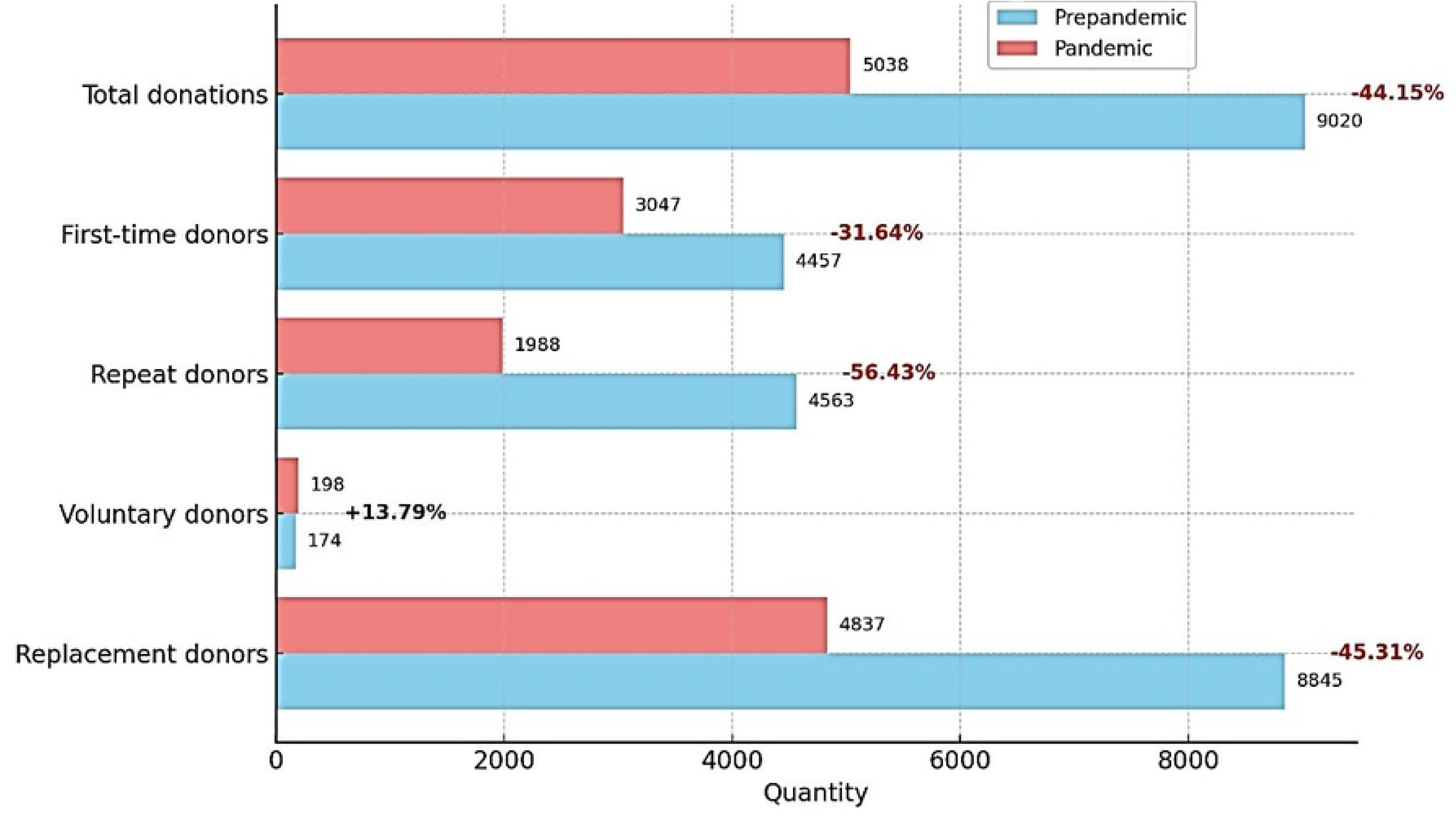
Comparison of total donations and donor types between the prepandemic and pandemic periods

\**Values in red and green indicate the percentage change between the two periods*

Similarly, a 38.5% reduction in the total number of transfusions was also recorded. Among the transfused blood components, there was a 41.62% decrease in whole blood, a 30.05% decrease in platelets, a 59.04% decrease in cryoprecipitate, and a 45.49% decrease in fresh frozen plasma. Regarding hospital transfusion requests, a 40.11% reduction was observed. By clinical area, the greatest decrease occurred in the surgical area, with a 59.22% decline (Fig 3).

**Fig 3.**
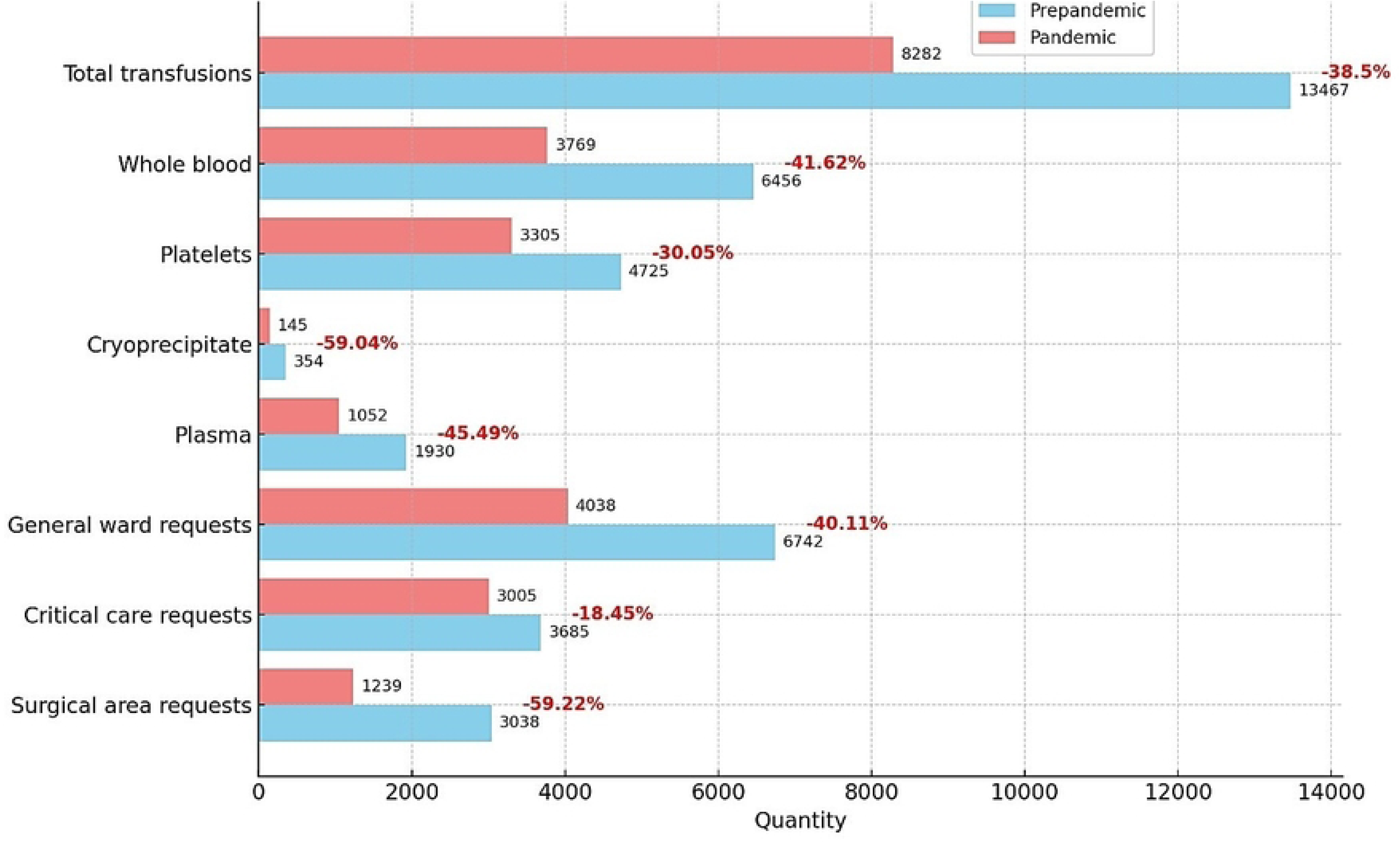
Comparison of transfusions and transfusion requests between the prepandemic and pandemic periods.

\**Values in red represent the percentage decrease between the two periods*.

The comparative analysis between the pre-pandemic and pandemic periods revealed statistically significant differences in the monthly donation characteristics. During the pandemic, the average monthly number of male donors decreased (173.93; 95% CI: 116.10– 231.75; p < 0.01), and the mean donor age was slightly lower (2.01; 95% CI: 1.44–2.57; p < 0.01). Regarding donation behavior, there was a significant reduction in the monthly number of donations during the pandemic (256.75; 95% CI: 167.46–346.04; p < 0.01). Similarly, significant decreases were observed in the number of replacement donors (259.71; 95% CI: 165.67–353.75; p < 0.01), repeat donors (173.01; 95% CI: 134.30–211.71; p < 0.01), and new donors (83.97; 95% CI: 15.68–152.27; p = 0.02). No statistically significant differences were found in the number of voluntary donors (p = 0.90) or in the number of seropositive donations (p = 0.47) between the two periods (Table 1).

**Table 1.** General characteristics of blood donations in relation to the COVID-19 pandemic.

|  | **Prepandemic**<br>**(Mean ± SD)**<br>**(n=9 020)** | **Pandemic**<br>**(Mean ± SD)**<br>**(n=5 038)** | **Difference** | **p** |
| --- | --- | --- | --- | --- |
| **Sociodemographic, month** |  |  |  |  |
| Number of male donors | 438.93 ± 49.46 | 265.00 ± 91.76 | 173.93 [116.10-231.75] | <0.01<sup>b</sup> |
| Average age | 38.12 ± 0.90 | 36.11 ± 0.40 | 2.01 [1.44-2.57] | <0.01<sup>a</sup> |
| **Donation, month** |  |  |  |  |
| Total donations | 644.29 ± 70.13 | 387.54 ± 145.14 | 256.75 [167.46-346.04] | <0.01<sup>b</sup> |
| Seropositive donations | 7.12 ± 1.89 | 7.58 ± 1.29 | -0.45 [-1.73-0.82] | 0.47<sup>c</sup> |
| Number of voluntary donors | 12.43 ± 21.45 | 15.23 ± 19.29 | -2.80 [-19.02-13.41] | 0.90<sup>a</sup> |
| Number of replacement donors | 631.79 ± 67.98 | 372.08 ± 146.48 | 259.71 [165.67-353.75] | <0.01<sup>c</sup> |
| Number of first time donors | 318.36 ± 66.15 | 234.38 ± 103.45 | 83.97 [15.68-152.27] | 0.02<sup>b</sup> |
| Number of repeat donors | 325.93 ± 48.50 | 152.92 ± 49.11 | 173.01 [134.30-211.71] | <0.01<sup>a</sup> |
| <sup>a</sup>= Calculated using Mann–Whitney U test, <sup>b</sup>= Calculated using Student’s t-test, <sup>c</sup>= Calculated using Student’s t-test with unequal variances. **X**: mean, **SD**: standard deviation, **n**: sample size<br>**\*Prepandemic period:** from January 1, 2019 to March 5, 2020<br>**\*Pandemic period:** from March 6, 2020 to March 6, 2021 |  |  |  |  |

In terms of the total number of monthly transfusions, a significant decrease was observed compared to the pre-pandemic period (324.85; 95% CI: 213.35–436.35, p < 0.01). Among the transfused blood components, there was a significant reduction in whole blood transfusions (171.22 units; 95% CI: 120.49–221.95, p < 0.01), platelets (83.27 units; 95% CI: 22.62–143.92, p < 0.01), and plasma (56.93 units; 95% CI: 31.95–81.91, p < 0.01). For cryoprecipitate, although a reduction was noted, it did not reach statistical significance (p = 0.07). As for transfusion requests by hospital area, a statistically significant decrease was observed in surgical units (121.69; 95% CI: 82.68–160.70, p < 0.01) and general hospitalization wards (170.96; 95% CI: 101.09–240.82, p < 0.01). In contrast, the decrease in requests from critical care units was not statistically significant (p = 0.27) (Table 2).

**Table 2.**
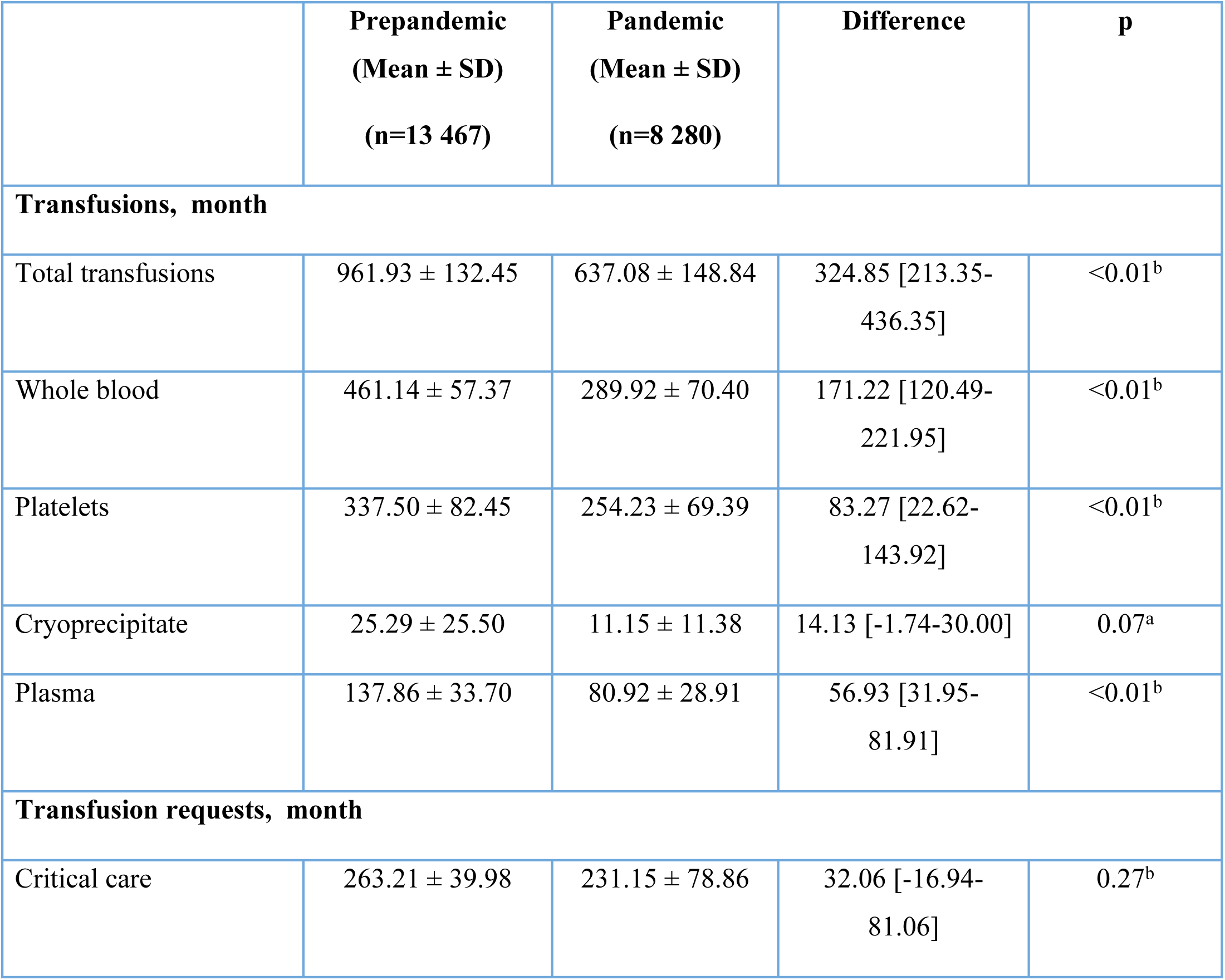
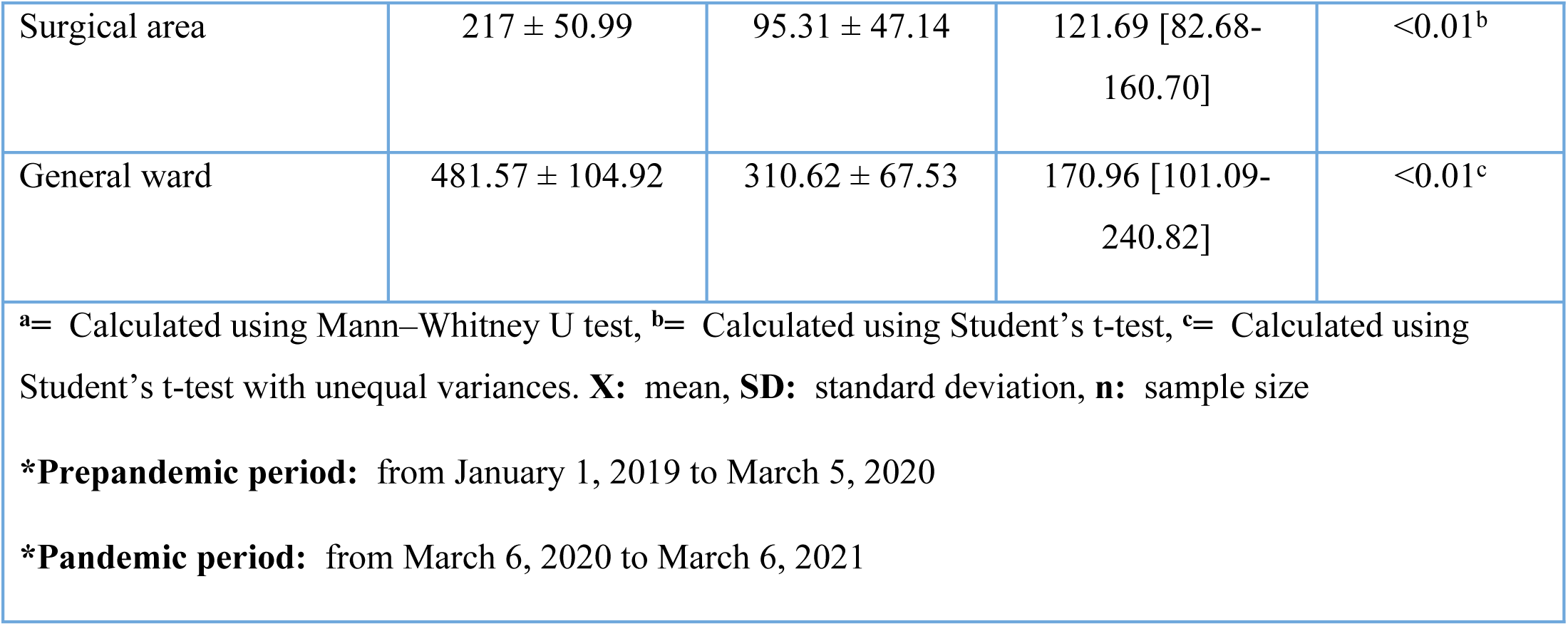
General characteristics of blood transfusions in relation to the COVID-19 pandemic.

|  |  |  |  |  |
| --- | --- | --- | --- | --- |
|  | **Prepandemic**<br>**(Mean ± SD)**<br>**(n=13 467)** | **Pandemic**<br>**(Mean ± SD)**<br>**(n=8 280)** | **Difference** | **p** |
| **Transfusions, month** |  |  |  |  |
| Total transfusions | 961.93 ± 132.45 | 637.08 ± 148.84 | 324.85 [213.35-436.35] | <0.01<sup>b</sup> |
| Whole blood | 461.14 ± 57.37 | 289.92 ± 70.40 | 171.22 [120.49-221.95] | <0.01<sup>b</sup> |
| Platelets | 337.50 ± 82.45 | 254.23 ± 69.39 | 83.27 [22.62-143.92] | <0.01<sup>b</sup> |
| Cryoprecipitate | 25.29 ± 25.50 | 11.15 ± 11.38 | 14.13 [-1.74-30.00] | 0.07<sup>a</sup> |
| Plasma | 137.86 ± 33.70 | 80.92 ± 28.91 | 56.93 [31.95-81.91] | <0.01<sup>b</sup> |
| **Transfusion requests, month** |  |  |  |  |
| Critical care | 263.21 ± 39.98 | 231.15 ± 78.86 | 32.06 [-16.94-81.06] | 0.27<sup>b</sup> |
| Surgical area | 217 ± 50.99 | 95.31 ± 47.14 | 121.69 [82.68-160.70] | <0.01<sup>b</sup> |
| General ward | 481.57 ± 104.92 | 310.62 ± 67.53 | 170.96 [101.09-240.82] | <0.01<sup>c</sup> |
| <sup>a</sup>= Calculated using Mann–Whitney U test, <sup>b</sup>= Calculated using Student's t-test, <sup>c</sup>= Calculated using Student's t-test with unequal variances. **X**: mean, **SD**: standard deviation, **n**: sample size<br>**\*Prepandemic period:** from January 1, 2019 to March 5, 2020<br>**\*Pandemic period:** from March 6, 2020 to March 6, 2021 |  |  |  |  |

## Discussion

he results of this study demonstrate a significant impact of a pandemic on the blood donation and transfusion system in a high-complexity national hospital. The comparative analysis between the pre-pandemic and pandemic periods revealed a monthly decrease of 256.75 donations and 324.85 transfusions, reflecting the disruption caused by the COVID-19 pandemic in the blood supply chain and the resulting imbalance between supply and demand. Studies conducted in Singapore have shown that public health emergencies lead to unpredictable fluctuations in blood availability, requiring adaptive strategies to mitigate their impact on the functioning of blood banks [1,3]. In China, it was reported that the rapid implementation of coordinated measures in blood centers helped minimize supply disruptions during the initial outbreak, underscoring the importance of organized responses in times of crisis [13].

The 44% reduction in total donations, including a notable 45.31% decrease in replacement donations, highlights the fragility of transfusion systems that rely heavily on this donation model, as is the case in Peru. Similar declines during the pandemic have been reported in countries such as Iran, Canada, Saudi Arabia, and China, with donation activity decreasing between 30% and 67% [5,7,9,14]. Although the present study recorded a 13.79% increase in voluntary donations, they continued to represent less than 5% of total donations, reflecting Peru’s historical dependence on replacement donations [4,15]. Furthermore, studies from Germany have shown that fear of infection, as well as reduced moral obligation and self-efficacy to donate, negatively affected blood donation intentions particularly among inactive or first-time donors. This issue may persist even beyond the acute phase of the health crisis [10).

Transfusion requests decreased by 40.11%, with an even more pronounced 59.22% reduction in surgical areas. This decline can be explained by hospital reorganization and the suspension of elective surgeries, measures implemented to prioritize COVID-19 care. Similar trends were reported in Spain and China [7,13]. The reduced demand translated into significant decreases in the transfusion of key blood components, including whole blood, cryoprecipitate, and fresh frozen plasma, reflecting the variability of demand and the challenges of maintaining a balanced supply during health emergencies [4,16]. Globally, it has been documented that the restructuring of medical services and the implementation of blood conservation and rational use strategies contributed to optimizing availability and reducing waste of blood products during the crisis [5].

In Peru, the high dependence on replacement donations, which account for over 90% of total donations, reveals a major vulnerability in the sustainability of the blood supply a situation similarly observed in other low and middle income countries [4,15]. Studies conducted in Cameroon and elsewhere have shown that a low proportion of voluntary donations poses a risk to transfusion safety and service continuity, making it essential to promote the recruitment and retention of regular donors [9,17]. The short shelf life of certain blood components particularly platelets, which last only five days further emphasizes the need for robust logistics systems and flexible planning to reduce the risk of shortages during emergencies [5,16]. Altogether, these findings underscore not only the vulnerability of replacement-dependent transfusion systems, but also the urgent need to transition toward a voluntary donation model that is more resilient to health crises.

The findings of this study emphasize the importance of developing dynamic and adaptive emergency plans to anticipate and manage abrupt fluctuations in blood donations and transfusions during health crises. Diversifying the donor base, strengthening voluntary donation, and optimizing logistics and inventory management are essential to ensure the resilience and sustainability of the blood supply under emergency conditions [3,5]. In addition, it is necessary to design effective communication strategies and educational campaigns, such as those implemented in China and Italy, which have proven crucial in mitigating donation declines and enhancing public trust in the safety of the donation process during the pandemic [3,5,13,18]. Furthermore, future multicenter studies could explore the sociodemographic determinants of voluntary donation and assess the impact of audiovisual and social media campaigns on donor behavior.

### Limitations

This study has several important limitations. First, its single-center design limits the generalizability of the findings to the national level. Additionally, the lack of key sociodemographic variables such as education level and income may have limited the analysis, as these factors could influence the frequency of donations and transfusions. Moreover, the analysis focused exclusively on the early waves of the pandemic, without accounting for the potential effects of vaccination rollout or the emergence of new viral variants. Nonetheless, the study’s methodological rigor and the quality of the data collected represent strengths that provide valuable evidence for health planning in similar contexts, contributing to the emerging body of literature in Latin America on the impact of the pandemic on transfusion services.

## Conclusions

The COVID-19 pandemic had a significant negative impact on the frequency and volume of blood donations and transfusions at the Blood Bank of Hospital Daniel Alcides Carrión, leading to a concerning imbalance in the availability of blood components. This situation reflects the vulnerability of transfusion systems that rely heavily on replacement donations, and highlights the urgent need to strengthen transfusion infrastructure and develop strategies for the recruitment and retention of voluntary donors.

This experience underscores the importance of implementing regional and local contingency plans to ensure the safety and adequacy of the blood supply during future health emergencies or natural disasters. Furthermore, it calls attention to the role of public education campaigns in combating misinformation and promoting safe blood donation practices during crises.

## Data Availability

All relevant data are within the manuscript and its Supporting Information files

## Notes

### Competing Interest Statement

The authors have declared no competing interest.

### Funding Statement

The author(s) received no specific funding for this work.

### Author Declarations

This was a non-interventional study based on an anonymized database. It was approved by the Ethics Committee of Universidad Científica del Sur under registration number 804-2021-PRE15, and it received the corresponding institutional authorization (Document No. 3350-2023-HNDAC) for access to the Blood Bank database.

